# Sensitivity, specificity and predictive values of molecular and serological tests for COVID-19: A longitudinal study in emergency room

**DOI:** 10.1101/2020.08.09.20171355

**Authors:** Zeno Bisoffi, Elena Pomari, Michela Deiana, Chiara Piubelli, Niccolò Ronzoni, Anna Beltrame, Giulia Bertoli, Niccolò Riccardi, Francesca Perandin, Fabio Formenti, Federico Gobbi, Dora Buonfrate, Ronaldo Silva

**Affiliations:** Department of Infectious, Tropical Diseases and Microbiology, IRCCS Sacro Cuore Don Calabria Hospital, Negrar di Valpolicella, Verona, Italy; Department of Diagnostics and Public Health, University of Verona, Verona, Italy

## Abstract

Accuracy of diagnostic tests is essential for suspected cases of Coronavirus Disease 2019 (COVID-19). This study aimed to assess the sensitivity, specificity and positive and negative predictive value (PPV and NPV) of molecular and serological tests for the diagnosis of SARS-CoV-2 infection. A total of 346 consenting, adult patients were enrolled at the emergency room of IRCCS Sacro Cuore Don Calabria Hospital, Negrar, Italy. We evaluated three RT-PCR methods including six different gene targets; five serologic rapid diagnostic tests (RDT); one ELISA test. The final classification of infected/not infected patients was performed using Latent Class Analysis in combination with clinical re-assessment of incongruous cases and was the basis for the main analysis of accuracy.

Of 346 patients consecutively enrolled, 85 (24.6%) were classified as infected. The molecular test with the highest sensitivity, specificity, PPV and NPV was RQ-SARS-nCoV-2 with 91.8% (C.I. 83.8-96.6), 100% (C.I. 98.6-100.0), 100.0% (C.I. 95.4-100.0) and 97.4% (C.I. 94.7-98.9) respectively, followed by CDC 2019-nCoV with 76.2% (C.I. 65.7-84.8), 99.6% (C.I. 97.9-100.0), 98.5% (C.I. 91.7-100.0) and 92.9% (C.I. 89.2-95.6) and by in-house test targeting *E-RdRp* with 61.2% (C.I. 50.0-71.6), 99.6% (C.I. 97.9-100.0), 98.1% (C.I. 89.9-100.0) and 88.7% (C.I. 84.6-92.1). The analyses on single gene targets found the highest sensitivity for *S* and *RdRp* of the RQ-SARS-nCoV-2 (both with sensitivity 94.1%, C.I. 86.8-98.1). The in-house *RdRp* had the lowest sensitivity (62.4%, C.I. 51.2-72.6). The specificity ranged from 99.2% (C.I. 97.3-99.9) for in-house *RdRp* and *N2* to 95.0% (C.I. 91.6-97.3) for E. The PPV ranged from 97.1% (C.I. 89.8-99.6) of *N2* to 85.4% (C.I. 76.3-92.00) of E, and the NPV from 98.1% (C.I. 95.5-99.4) of gene *S* to 89.0% (C.I. 84.8-92.4) of in-house *RdRp*. All serological tests had <50% sensitivity and low PPV and NPV. One RDT (VivaDiag IgM) had high specificity (98.5%, with PPV 84.0%), but poor sensitivity (24.7%). Molecular tests for SARS-CoV-2 infection showed excellent specificity, but significant differences in sensitivity. As expected, serological tests have limited utility in a clinical context.

## INTRODUCTION

As of today (17^th^ July 2020), the Severe Acute Respiratory Syndrome Coronavirus 2 (SARS-CoV-2) has infected 13 616 593 individuals, caused 585 727 deaths and has spread to virtually all countries^1^. Italy has been the first affected country in Europe and one of the most affected worldwide. The diagnosis of SARS-CoV-2 infection is based on standardized molecular methods, usually performed on nasal/pharyngeal swabs^2^. However, the accuracy of the different methods has yet to be properly assessed. Sensitivity for instance depends on the method itself^3^, the correct execution of the nasal and pharyngeal swab^4^, and also the timing since exposure and onset of symptoms ^5-7^ False negative results may cause mismanagement and nosocomial or community transmission^8,9^. False positive results imply the risk for a patient suffering from another condition to be erroneously admitted to a Coronavirus Disease (COVID) unit, or quarantined at home, besides triggering a complex but useless contact tracing^10^.

Many antibody-based tests, including rapid diagnostic tests (RDT) have been developed ^11-14^ and marketed and some have already been evaluated in retrospective studies^15,16^. Most RDT can be performed by simple finger prick and the result is available in a brief delay. However, the delay between onset of symptoms and detectability of antibodies obviously hampers the sensitivity of RDTs in case of recent infections, thus their diagnostic value at symptoms onset is disputable^17-22^. The main use of all serologic tests is now restricted to screening and epidemiologic purposes. It has been suggested, however, that an RDT in combination with RT-PCR might be useful in clinical practice, too; however, no supporting data have been provided as yet to this hypothesis ^3^.

### Study objectives

The primary objective of this study was to assess sensitivity, specificity and predictive values of three widely used Reverse Trascriptase-real time PCR (RT-PCR), with six different gene targets, for diagnosis of COVID-19 in clinically suspect cases. The secondary objective was to define whether any of six serological tests, five IgG-IgM rapid diagnostic tests (RDT) and an ELISA IgA-IgG test, may be of diagnostic usefulness.

## METHODS

This paper refers to STARD guidelines^23^ for the reporting of diagnostic tests accuracy. The assessment was carried out using the statistical technique of Latent Class Analysis.

### Type of study

Observational, prospective diagnostic study. Data collection was planned before performing the index tests and the reference standard tests.

The study was performed at IRCCS Sacro Cuore Don Calabria Hospital, Negrar, a reference Institute for Infectious and Tropical Diseases in Italy. Patients were enrolled at the first diagnostic workout at the emergency room (ER).

### Study cohort and participant recruitment

All consecutive patients presenting to the ER with clinical suspicion of COVID-19 and submitted to diagnostic tests for SARS-CoV-2 were eligible. The essential clinical and laboratory data were recorded in an electronic Case Report Form (e-CRF). Enrolment continued until completion of the required sample size.

#### Inclusion criteria

Adult male and female patients. Consent to participate to the study and to the donation of biological samples.

#### Exclusion criteria

Missing or inadequate samples.

Test methods

#### Index tests

All the index tests were performed on samples consecutively collected and stored at −80 °C: nasal/pharyngeal swabs for molecular tests and serum for serological methods.

a. Molecular tests (RT-PCR)

1. RealQuality RQ-SARS-nCoV-2 assay (cod. RQ-130, AB Analitica, Italy), targeting the *spike protein* gene *(S)* and the *RNA-dependent RNA polymerase* gene *(RdRp)*.
2. CDC 2019-Novel Coronavirus (2019-nCoV) Real-Time RT-PCR Diagnostic Panel, targeting the *nucleocapsidprotein* gene (N), regions *N1* and *N2*.
3. In-house RT-PCR protocol performed on nasal/pharyngeal swabs, targeting the *envelope protein* gene *(E)* in the first-line screening assay, followed by confirmatory testing with the *RdRp* gene (same gene as in RQ-130, AB Analitica, but with different molecular targets)^24^.
b. IgG/IgM immune chromatographic RDT

4. 2019-nCoV IgG/IgM Rapid Test Cassette (JusCheck, Acro Biotech, USA)
5. COVID-19 IgG/IgM Rapid Test Cassette (Femometer Hangzhou Clongene Biotech, China)
6. COVID-19 IgG/IgM Rapid Test (Prima Professional, Switzerland)
7. VivaDiag 2019-nCoV IgG/IgM rapidTest (VivaCheck Biotech, China)
c. IgG/IgM immunofluorescence RDT

8. DiaGreat 2019-nCoV IgG/IgM antibody Determination Kit (Nuclear Laser Medicine, Italy).
d. IgG/IgA ELISA test

9. Anti-SARS-CoV-2 ELISA IgA/IgG (Euroimmun, Germany).

The test methods are described in detail in the supplementary appendix.

#### Blinding

Each test was executed by experienced lab personnel of the reference laboratory independently. The lab professionals were not aware of the clinical data of the subjects and did not know in advance the results of any other test.

#### Reference Standard based on Latent Class Analysis (LCA)

LCA is the preferred method for evaluating a diagnostic test in the absence of a gold standard^25-29^ Typically for SARS-CoV-2 infection, no single test can be considered as a gold standard. Tests based on RT-PCR are highly specific, but their sensitivity may not be optimal. The LCA method is summarized in the paragraph Statistical methods and analysis.

#### Evaluation criteria of molecular tests accuracy

Each of the three molecular tests is based on two gene targets, both of which are required to be positive in order to diagnose the infection ^24^ (https://www.fda.gov/media/134922/download). A first, restrictive analysis was performed based on this criterion, indeterminate results (only one positive gene target) being classified as negative. However, a further analysis of accuracy was also performed on single gene targets.

### Statistical methods and analysis

Sample size. The sample size calculation was based on the desired 10% width of the 95% exact confidence intervals around the point estimates and on a minimal acceptable sensitivity of 95%, and identical specificity. To assure an adequate sample size, 94 patients with SARS-CoV-2 infection and as many negatives were needed. Based on figures observed in the weeks before study onset, since we expected a proportion of infections of about 25% and we estimated an enrollment of 376 subjects. We planned to enroll 400 subjects to account for possible altered or invalid samples. Demographic and clinical data were summarized using descriptive statistics and measures of variability and precision. All parameters were reported with 95% confidence intervals (CI). For proportions the exact Clopper-Pearson CI was computed.

Diagnostic tests results were presented in contingency tables where patient’s disease status was inferred based on probabilistic models using LCA. This is a statistical method used to classify unobserved groups in a population based on a chosen set of indicators. In LCA it is assumed that the true patient’s condition is unknown, i.e, is a latent class ^29^, which can be related to a set of diagnostic test results, clinical and paraclinical variables, through latent class models (LCM). Each class corresponds to a possible condition of the patient, thus a 2-class model will classify patients as presumably having/not having the condition, while a 3-class model will identify a third group of patients of uncertain classification. The best model and number of classes are chosen based on appropriate statistic methods such as Akaike’s information criterion (AIC) or likelihood ratio test. Computerized medical records of patients with uncertain diagnosis according to LCA were reviewed (including all tests repeated in the following days, if any) in order to obtain a reasonably certain diagnosis. Sensitivities, specificities and predictive values of the index tests were calculated based on this final diagnosis.

Data analysis were performed using SAS software, version 9.4 (SAS Institute, Inc., Cary, NC, USA). Statistical significance level was fixed at 0.05. LCMs were built using the LCA procedure with parameters estimated by maximum likelihood using the Expectation-maximization algorithm. A rho prior of strength 1 was used when needed, to avoid estimations on the boundary of the parameters space. Missing values on any diagnostic tests were handled by the LCM. Standard procedures were used for the verification of the assumption of conditional independence between diagnostics tests. To reproduce results, use seed=1979 in proc LCA.

Further details on the method and on the models tested are available in the Appendix.

Composite Reference Standard (CRS). This is an alternative method for assessing test accuracy when a gold standard is missing. Exploratory analyses were also carried out using CRS for a classification of the study subjects. They are also reported in the Appendix.

### Ethical clearance

The study protocol was approved by the pertinent Ethical Committee (Comitato Etico per la Sperimentazione clinica delle Province di Verona e Rovigo, Protocol N. 19408, 2nd April 2020 and following amendment, protocol N. 33102, approved on 10th June 2020). All the patients included gave their consent to the storage of biological samples in the “Tropica Biobank” and use of related results for research purposes, as per routine procedure in our hospital.

### Protocol registration

The protocol has been registered at http://www.isrctn.com/ with the ID ISRCTN13990999.

## RESULTS

The study was carried out from 1th March to 9th May 2020. Three hundred and forty-six patients were consecutively enrolled (Study Flow Chart, Figure 1). Their main demographic, clinical, laboratory and imaging characteristics are summarized in Figure 2.

**Figure 1.**
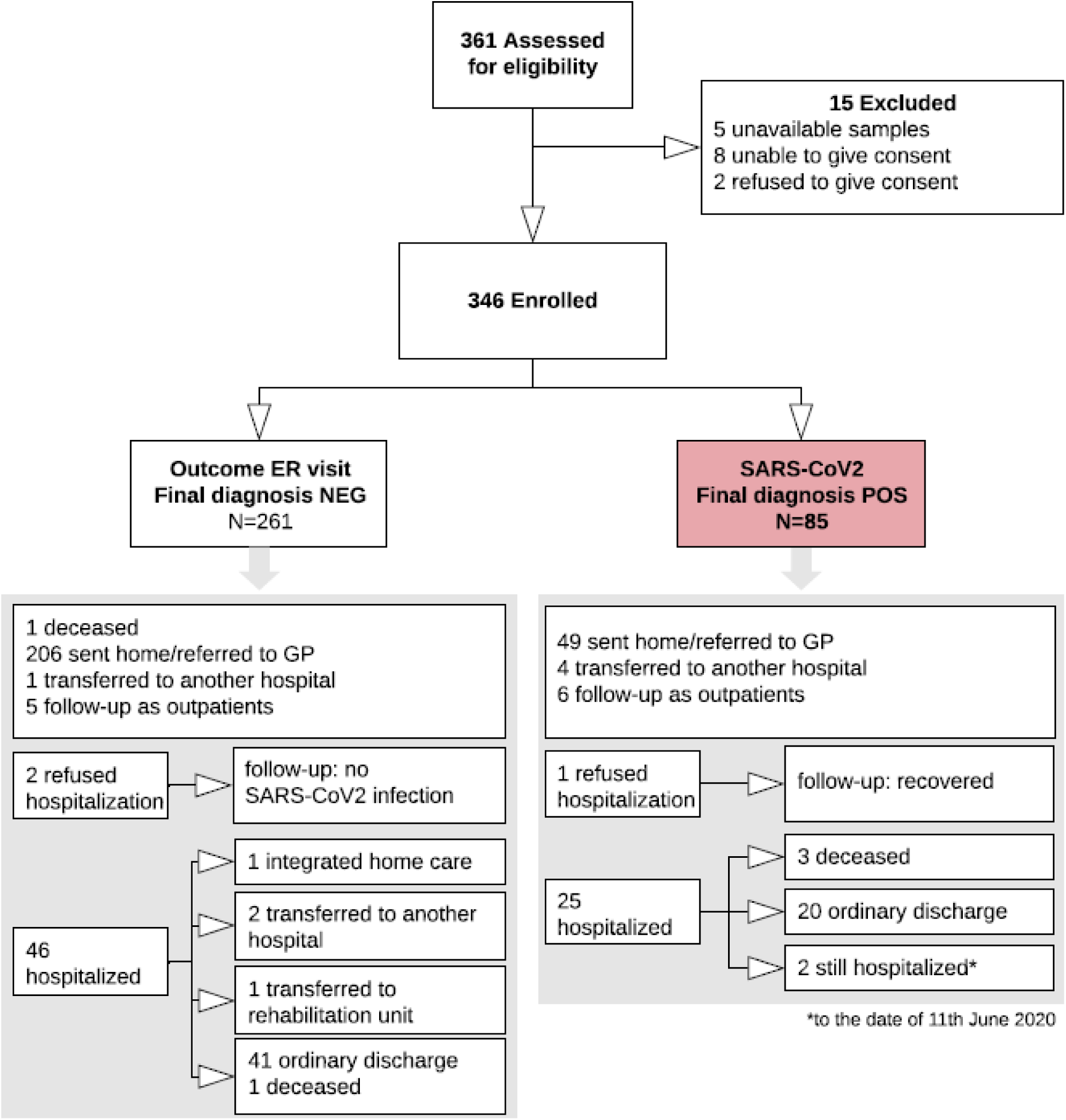
Study Flow chart.

**Figure 2.**
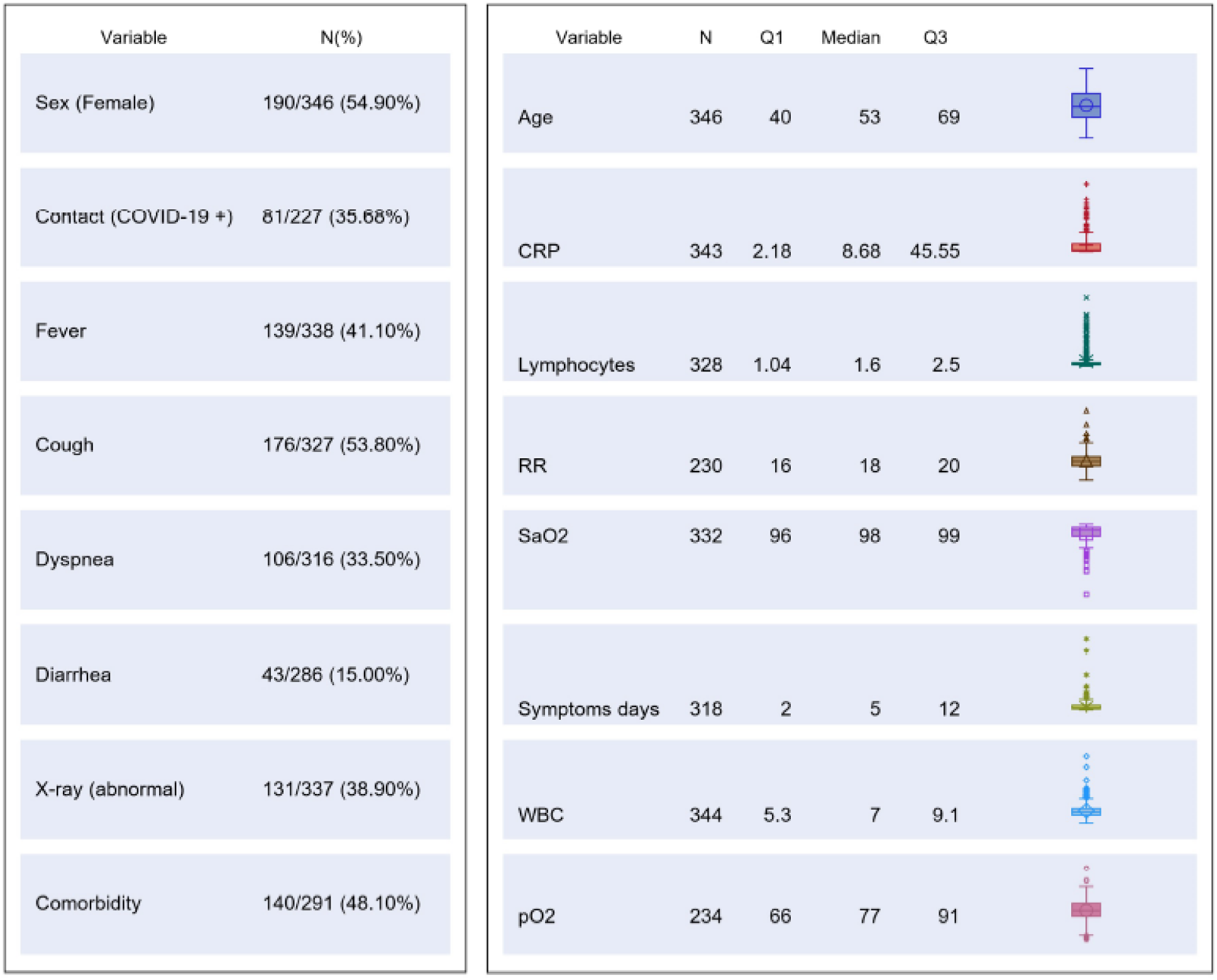
Table chart on main demographic, clinical, laboratory and imaging characteristics of the study population.

Clinical management was based on the result of the molecular test used at ER.

### Latent Class Analysis (LCA)

Based on the best fitted LCA model (with three LCA classes) 332/346 patients (96%) could be classified as infected or non infected with virtual certainty. Fourteen subjects (4%) could not be attributed with certainty to the infected or uninfected group. The computerized medical records of the latter patients were reviewed (including all tests repeated in the following days, if any) in order to reach a reasonably certain diagnosis. Three patients for whom the final diagnosis remained doubtful after reassessment were tested with an additional ELISA Anti-SARS-CoV-2 IgG test (Abbott), performed 6 to 8 weeks after the first diagnosis. The three of them tested IgG negative and were then finally classified as non-infected. Furthermore, an assessment of all medical records of patients with at least one discordant gene target result was performed. Finally, 85 out of 346 patients (24.6%) were classified as infected, and 261 (75.4%) as non-infected. Based on these denominators and applying the restrictive criterion (both gene targets required to be positive to define a case of infection), the test accuracy results are summarized in Figure 3. The molecular test with the highest sensitivity was RQ-SARS-nCoV-2 (91.8%, C.I. 83.8-96.6), followed by CDC 2019-nCoV (76.2%, C.I. 65.7-84.8) and by in-house primary reference test targeting *E-RdRp* (61.2%, C.I. 50.0-71.6). The specificity was 100% for RQ-SARS-nCoV-2 (C.I. 98.6-100.0) and 99.6% for the other two tests (C.I. 97.9-100.0). The test with the highest PPV and NPV was, again, RQ-SARS-nCoV-2 (100.0%, C.I. 95.4-100.0 and 97.4%, C.I. 94.7-98.9, respectively), followed by CDC 2019-nCoV (98.5%, C.I. 91.7-100.0 and 92.9%, C.I. 89.2-95.6) and by *E-RdRp* test (98.1%, C.I. 89.9-100.0 and 88.7%, C.I. 84.6-92.1).

**Figure 3.**
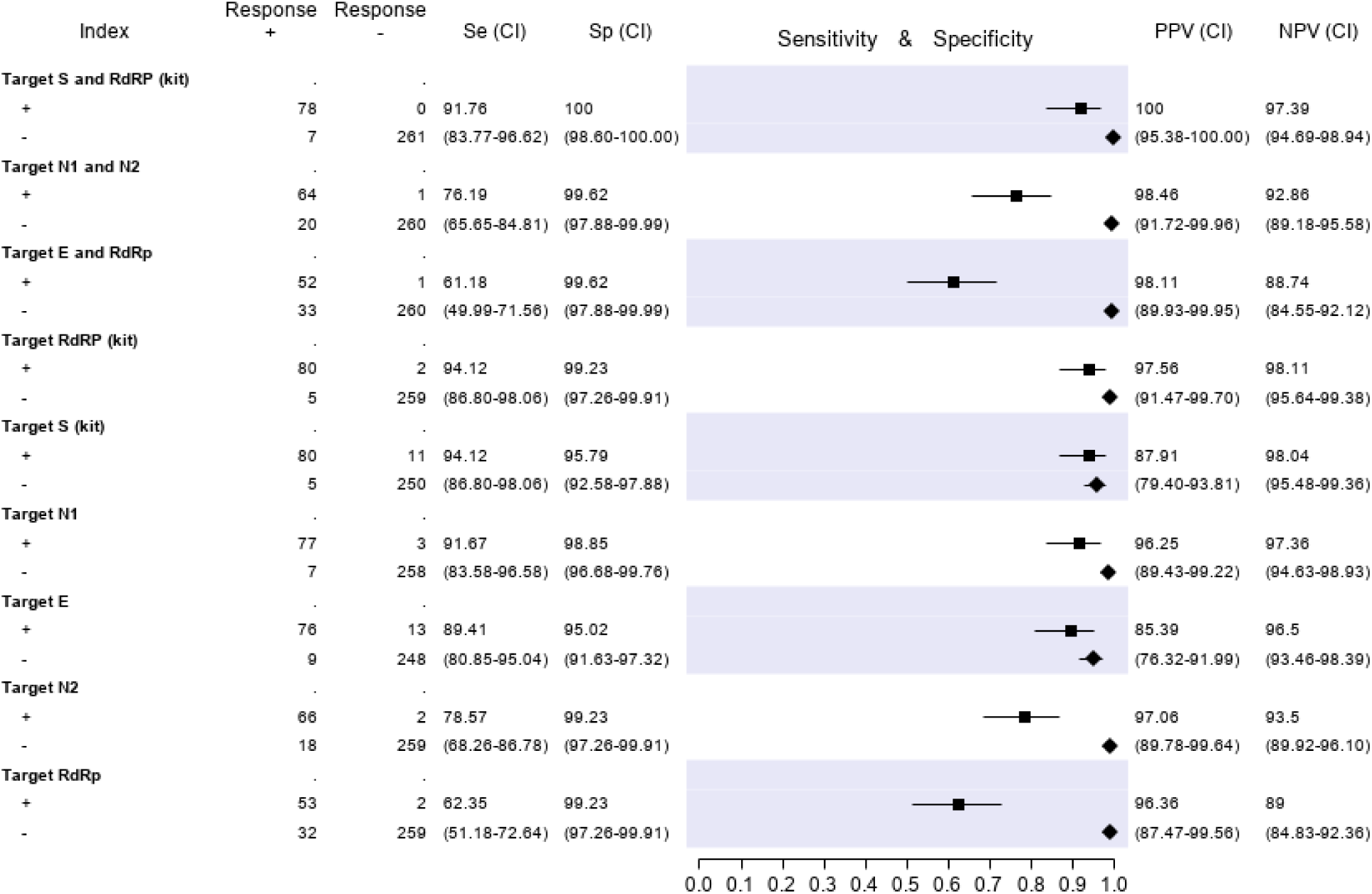
Sensitivity (filled squares), specificity (filled rhombus) and predictive values of 3 molecular tests and b) of single gene targets according to the final classification (Infected =85, not infective=261) Target S and RdRp (kit)= RealQuality RQ-SARS-nCoV-2 assay (cod. RQ-130, AB Analitica, Italy), targeting the *spike protein* gene (*S*) and the *RNA-dependent RNA polymerase* gene (*RdRp*). Target N1 and N2 = CDC 2019-Novel Coronavirus (2019-nCoV) Real-Time RT-PCR Diagnostic Panel, targeting the *nucleocapsid protein* gene (*N*), regions *N1* and *N2*. Target E and RdRp = *in-house* RT-PCR protocol, targeting the *envelope protein* gene (*E*) and the *RdRp*.

The results of analyses on single gene targets are reported in Figure 3. The assays with the highest sensitivity were those targeting *S* and *RdRp* of the RQ-SARS-nCoV-2 (both with sensitivity 94.1%, C.I. 86.8-98.1). The in-house *RdRp* was the target with the lowest sensitivity (62.4%, C.I. 51.2-72.6). The specificity ranged from 99.2% (C.I. 97.3-99.9) for in-house *RdRp* and *N2* to 95.0% (C.I. 91.6-97.3) for gene E. The PPV ranged from 97.1% (C.I. 89.8-99.6) of *N2* gene to 85.4% (C.I. 76.3-92.00) of *E* gene, and the NPV from 98.1% (C.I. 95.5-99.4) of gene *S* to 89.0% (C.I. 84.8-92.4) of in-house *RdRp*.

### Concordance among the six gene targets

For 42 of the 85 subjects with positive final diagnosis (49%), all gene target results were concordant positive, while for 234 of the 261 subjects with positive final diagnosis (90%) they were concordant negative. The 70 records with at least one discordant result are represented graphically in Figure 4.

**Figure 4.**
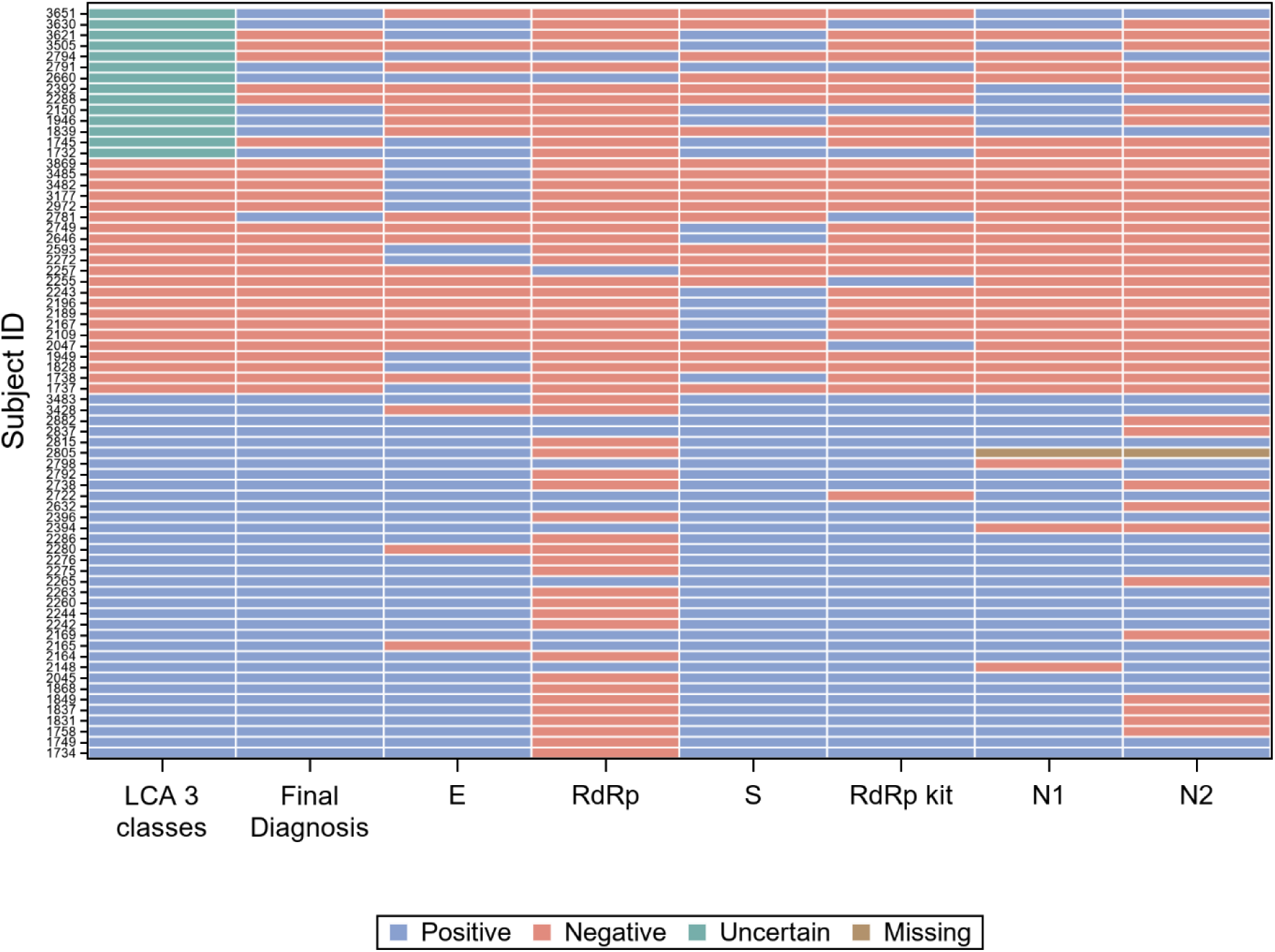
Visual representation of the 70 records with at least one discordant gene target result.

The results of the serological tests are reported in Figure 5. Briefly, the sensitivity ranged from 45.9% (Prima Professional IgM) to 21.2% (ELISA IgG); the specificity, from 98.5% (VivaDiag IgM) to 79.7% (Prima Professional IgM); the PPV, from 84.0% (VivaDiag IgM) to 44.1% (ELISA IgA); the NPV, from 82.3 (JusCheck IgM) to 79.1% (ELISA IgG).

**Figure 5.**
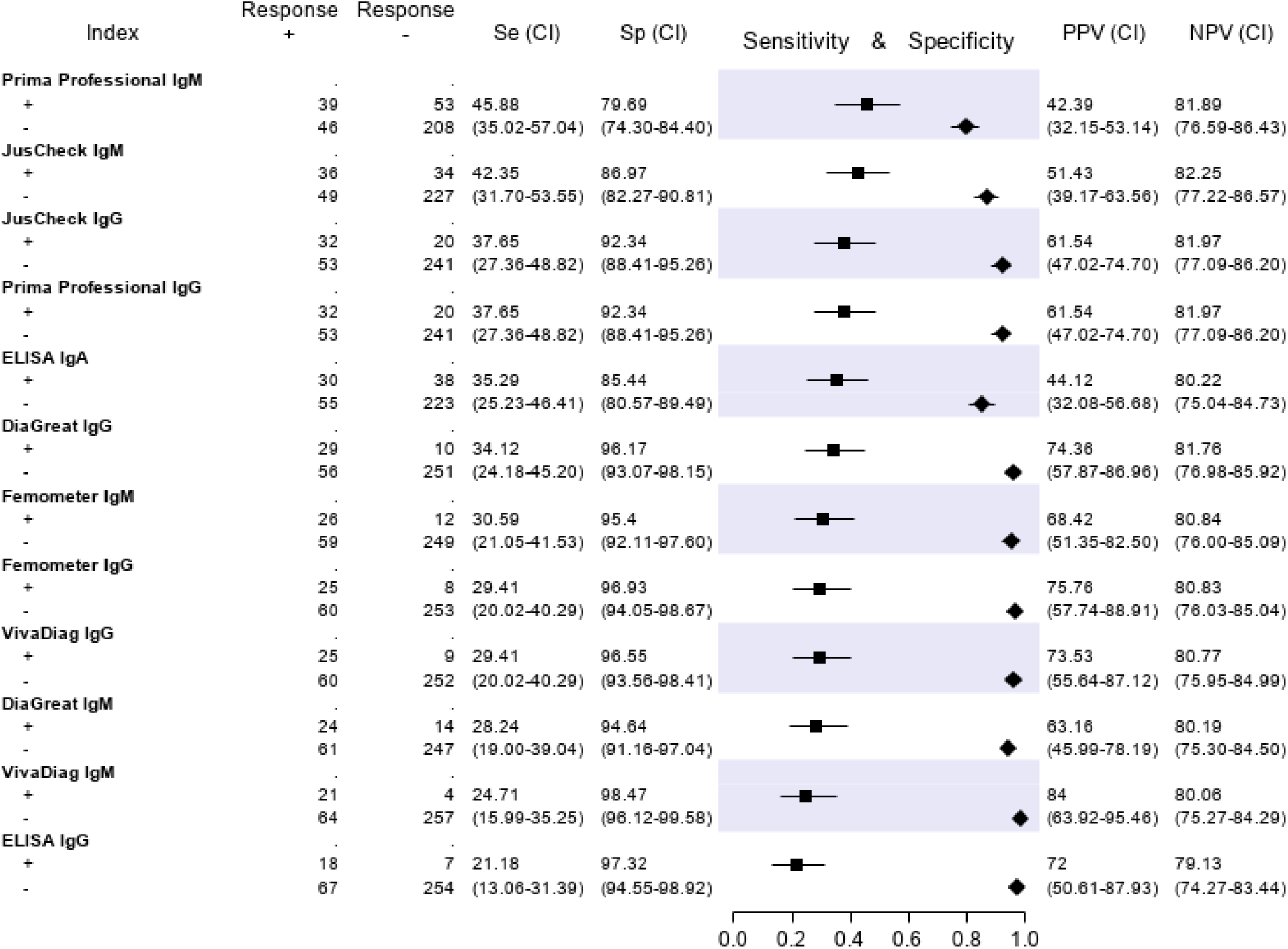
Sensitivity (filled squares), specificity (filled rhombus) and predictive values of 6 antibody tests according to the final classification (Infected =85, not infective=261)

## DISCUSSION

This is to our knowledge, the first formal longitudinal accuracy study of both molecular and serological tests for the diagnosis of SARS-COV-2 infection in suspected COVID-19 patients. The molecular tests showed significant differences in sensitivity, which was >90% only for RQ-SARS-nCoV-2. This raises concern on the current protocols for COVID-19 diagnosis, as most require at least two different SARS-CoV-2 genomic regions to be concordantly positive in order to classify a subject as infected^3,4,24,30-32^. Actually, early in the course of the epidemic, we realized that a number of patients who resulted positive at the first line screening test (E target), but negative at the confirmatory test *(RdRp* target), and who would have been then classified as non infected with SARS-CoV2, were most probably true positives, and we started managing patients accordingly. Interestingly, in accordance with our practice, current US FDA recommendations consider a single positive gene target as sufficient to validate the performance of RT-PCR assays for the diagnosis of COVID-19 (https://www.fda.gov/medical-devices/emergency-situations-medical-devices/faqs-testing-sars-cov-2).

However, the concordance among molecular gene targets was far from perfect. Figure 4 resumes the results of all single gene targets, compared with the patient classification according to LCA and with the final diagnosis, on the 70 samples (20% of the total) with at least one discordant result. It is immediately apparent from Figure 4 that the gene target *RdRp* of the in-house protocol (“RdRp” in the figure) accounts for a large proportion of the discordant results with the final diagnosis, while the same gene (with different molecular targets) of RealQuality RQ-SARS-nCoV-2 assay (“RdRp kit” in the figure) was the single gene that minimized discordant results.

Clearly however, whatever the test used, a variable proportion of truly infected patients may be missed, and patients with a high clinical suspicion should be carefully considered, even after a negative test result. Looking back at our study population, we realized that in fact, three out of five patients who had been initially wrongly diagnosed as negative, were in fact rightly managed as COVID-19 patients, due to a the clinical suspicion and consequent repetition of the test in subsequent nasal/pharyngeal swabs. However, two COVID-19 patients were incorrectly diagnosed and managed, as the infection was only demonstrated in retrospect.

The specificity was very high, expectedly, for all molecular tests when using the restrictive diagnostic criterion. When using the “relaxed” criterion of relying on a single gene target, the increased sensitivity is unsurprisingly mirrored by some loss in specificity. When dealing with clinically suspect cases in a phase of intense transmission, a high sensitivity is required, as missing a case would have serious consequences^10^. Also, in the presence of high clinical suspicion or pre-test probability, the positive predictive value of a test is obviously higher than in a screening context. Thus, recommendations on the correct interpretation of test results should be tailored to the clinical and epidemiological context. When the tests are used on suspect cases of COVID-19 in a phase of intense transmission, using the relaxed criterion is amply justified. However, when the same tests are used for screening purpose in a phase of low/very low viral circulation, relying on a “single-gene” approach would result in a higher proportion of false positive results.

No serological test showed an acceptable sensitivity nor specificity, confirming previous reports claiming that serological tests are unsuitable for clinical use in acutely ill patients and that their deployment should be limited to epidemiologic purposes^17,33-36^.

### Strengths

The study was conducted closely adhering to STARD guidelines. Moreover, in order to cope with the lack of a gold standard, the main analyses were carried out using LCA, upon condition that the chosen models well fitted the data. This study on a comparatively large cohort of patients suggests possible alternatives to current diagnostic protocols, in order to avoid the potentially dangerous, premature exclusion of a case of infection.

### Limitations

The sample size was slightly lower than the calculated number of 376 patients, due to the sharply decreasing number of new cases in the last period of the investigation. However, samples from almost all patients recruited were subsequently analyzed as there were no altered or invalid specimens, reaching a final final number of 346 patients, which was close to the planned sample size.

Despite the longitudinal study design, some clinical data were missing for a number of patients, which also reflects the inherent difficulties in performing clinical studies in emergency situations. However, for most variables included in the model the data set was sufficiently complete.

## CONCLUSION

The molecular tests evaluated here demonstrated significant differences in sensitivity. Accepting positive results in any single gene target for molecular diagnostics appears justified for cases with clinical suspicion of COVID-19 in an ER. Conversely, a confirmation of the diagnosis, based on positivity of multiple genomic regions, might be more appropriate when the test is deployed for screening purpose in a phase of low/very low viral circulation.

The serological tests included in this study did not demonstrate suitable for clinical use in acutely ill patients.

## Data Availability

All data generated or analysed during this study are included in this published article (and its supplementary information files).

## Acknowledgments

We thank the participating investigators from the IRCSS Sacro Cuore Don Calabria Hospital (Italy): Stefano Tais, Monica Degani, Marco Prato, Flavio Stefanini, Arjan Qefalia, Flavio Coato, Francesca Tamarozzi, Antonio Mori, Fabio Chesini, Giulia La Marca and Barbara Pajola.

